# Strengthening Deep-learning Models for Intracranial Hemorrhage Detection: Strongly Annotated Computed Tomography Images and Model Ensembles

**DOI:** 10.1101/2023.08.24.23293394

**Authors:** Dong-Wan Kang, Gi-Hun Park, Wi-Sun Ryu, Dawid Schellingerhout, Museong Kim, Yong Soo Kim, Chan-Young Park, Keon-Joo Lee, Moon-Ku Han, Han-Gil Jeong, Dong-Eog Kim

## Abstract

Multiple attempts at intracranial hemorrhage (ICH) detection using deep-learning techniques have been made and plagued with clinical failures. Most studies for ICH detection have insufficient data or weak annotations. We sought to determine whether a deep-learning algorithm for ICH detection trained on a strongly annotated dataset outperforms that trained on a weakly annotated dataset, and whether a weighted ensemble model that integrates separate models trained using datasets with different ICH subtypes is more accurate. We used publicly available brain CT scans from the Radiological Society of North America (27,861 CT scans, 3,528 ICHs) and AI-Hub (53,045 CT scans, 7,013 ICHs) for training datasets. For external testing, 600 CT scans (327 with ICH) from Dongguk University Medical Center and 386 CT scans (160 with ICH) from Qure.ai were used. DenseNet121, InceptionResNetV2, MobileNetV2, and VGG19 were trained on strongly and weakly annotated datasets and compared. We then developed a weighted ensemble model combining separate models trained on all ICH, subdural hemorrhage (SDH), subarachnoid hemorrhage (SAH), and small-lesion ICH cases. The final weighted ensemble model was compared to four well-known deep-learning models. Six neurologists reviewed difficult ICH cases after external testing. InceptionResNetV2, MobileNetV2, and VGG19 models outperformed when trained on strongly annotated datasets. A weighted ensemble model combining models trained on SDH, SAH, and small-lesion ICH had a higher AUC than a model only trained on all ICH cases. This model outperformed four well-known deep-learning models in terms of sensitivity, specificity, and AUC. Strongly annotated data are superior to weakly annotated data for training deep-learning algorithms. Since no model can capture all aspects of a complex task well, we developed a weighted ensemble model for ICH detection after training with large-scale strongly annotated CT scans. We also showed that a better understanding and management of cases challenging for AI and human is required to facilitate clinical use of ICH detection algorithms.

**Key Points:** **Question** Can a weighted ensemble method and strongly annotated training datasets develop a deep-learning model with high accuracy to detect intracranial hemorrhage?

**Findings** A deep-learning algorithm for detecting ICH trained with a strongly annotated dataset outperformed models trained with a weakly annotated dataset. After ensembling separate models that were trained with only SDH, SAH, and small-lesion ICH, a weighted ensemble model had a higher AUC.

**Meaning** This study suggests that to enhance the performance of deep-learning models, researchers should consider the distinct imaging characteristics of each hemorrhage subtype and use strongly annotated training datasets.

## Introduction

Intracranial hemorrhage (ICH) occurs in the intracranial space and encompasses the following six types: epidural hemorrhage (EDH), subdural hemorrhage (SDH), subarachnoid hemorrhage (SAH), intraparenchymal hemorrhage (IPH), intraventricular hemorrhage (IVH), and mixed hemorrhage. A timely and accurate diagnosis of ICH and its subtypes is critical for treatment, because of the high mortality and morbidity. In addition, assessing the location and extent of ICH is important for outcome prediction. However, neuroradiology training requires a significant investment of time and resources; accordingly, neuroradiologists are scarce in many countries.^1^ Without neuroradiologists’ assistance, doctors who see ICH patients often misdiagnose.^2^

Deep-learning algorithms have recently made progress in accurately detecting ICH on CT scans.^3^ However, their clinical use is limited due to challenges in identifying SDH or SAH.^4^ In contrast to other ICH subtypes, SDH is more likely to present in the subacute stage and has reduced CT attenuation, similar to that of brain tissue. SAH may also appear isoattenuating if there is only a small amount of blood mixed with cerebrospinal fluid.^5^ Moreover, a pseudo-SAH is not an uncommon finding; it manifests as high-attenuation areas along the basal cisterns, Sylvian fissure, tentorium cerebelli, or cortical sulci. These point out the importance of lesion location information for a more accurate diagnosis of ICH, particularly when the hemorrhage is difficult to detect due to its tiny volume or faint attenuation.

One of the challenges to increasing the accuracy of ICH detection algorithms is the lack of a large dataset with expert annotations, which would take a lot of effort and resources to produce.^6^ Thus, insufficient data or weak annotations have been employed in the majority of published research;^3,7,8^ In the weakly-supervised settings such as classification-based deep learning using images with annotations that are relatively easy to obtain (presence vs. absence of ICH), saliency maps could not locate the exact location of lesions.^9^

In this study, we hypothesized that a deep-learning model performs better if it is trained on a large annotated dataset with slice-wise manual segmentation, compared to the one trained on a weakly annotated dataset. To improve performance and robustness across all ICH subtypes and sizes, we also designed a weighted ensemble model to integrate multiple models trained on distinct strongly annotated datasets reflecting ICH features to minimize the prediction errors of each individual model. In addition, six experts reviewed challenging ICH cases after external testing of the final model.

## Methods

### 1. Model development with weakly and strongly annotated datasets

#### 1.1. Datasets

**Weakly annotated dataset** We used open data from the Radiological Society of North America (RSNA) comprising 27,861 brain CT scans (3,528 hemorrhages). Per slice, the neuroradiologists labeled the presence/absence of a hemorrhage without spatial annotation.^7^ **Strongly annotated dataset** We used 53,045 brain CT scans (7,013 with and 46,032 without ICH) from the AI-Hub directed by the Korean National Information Society Agency (https://aihub.or.kr/aidata/34101). The AI-Hub dataset was collected from six Korean university hospitals in 2020 as part of a large-scale data collection initiative for cerebrovascular disease. Each hospital’s neuroradiologist interpreted the CT scans, labeled the presence of hemorrhage per slice, and manually segmented the outline of the hemorrhage. A total of 7,013 CT scans with hemorrhages included 2,424 SAHs, 2,738 SDHs, 371 EDHs, 1,266 IVHs, 3,367 IPHs, and 2,833 mixed hemorrhage.

#### 1.2. Training and validation dataset

To compare the performance of the deep-learning models trained on weakly and strongly annotated datasets, and to account for different data sizes, we randomly selected the same number of slices with and without hemorrhage (n = 6,500 each) from the RSNA and AI-Hub datasets. For CT scans with hemorrhage, the same number of slices as those of IPH, IVH, SDH, EDH, and SAH (n = 1,300) were included, i.e. the hemorrhagic types were balanced. All training dataset images were pre-processed into four-channel input data (eMethods and eFigure 1).

#### 1.3. External test dataset

Two datasets were used for the external testing of the deep-learning models. The first dataset comprised 600 brain CT scans (327 with and 273 without ICH) from a tertiary hospital in Korea (Dongguk University Medical Center (DUMC)). The second was an open dataset (Qure.ai) comprising 386 CTs (160 with and 226 without ICH). A vascular neurologist and neuroradiologist performed consensus labeling for the DUMC dataset, and three neuroradiologists labeled the Qure.ai dataset using a majority vote. This study was approved by the institutional review board of DUMC and JLK Inc. (No. DUIH 2018-03-018 and 20220407-01).

#### 1.4. Comparison of models trained with weakly and strongly annotated datasets

DenseNet121,^10^ InceptionResNetV2,^11^ MobileNetV2,^12^ and VGG19^13^ were used for model development to compare deep-learning models trained on datasets with weak and strong annotations. For models using weakly annotated datasets, we used slice-wise hemorrhage labeling for both the RSNA and AI-Hub datasets (eFigure 2A-B). For classification loss, we compared the slice-wise model output and ground-truth labeling. The same input image was fed into the deep-learning model trained on the strongly annotated AI-Hub dataset (eFigure 2C). We extracted the saliency map from the last convolutional layer of each deep-learning model, compared it to the ground-truth hemorrhage segmentation, and computed the segmentation loss in addition to the classification loss to train the hemorrhage location.

We tested each model on an external test dataset and calculated sensitivity, specificity, and AUC and the threshold of 0.5 as the model’s performance. We used 500 bootstrap replications to calculate 95% confidence intervals. We used the DeLong test for AUC comparison.^14^

### 2. Ensemble model

#### 2.1. Training and test dataset

Considering class imbalance, we randomly selected an equal number of brain CT scans with and without ICH from the AI-Hub dataset (6,963 each). We trained five U-net based segmentation models (eFigure 3): Lesion segmentation model using all training datasets (Model 1), lesion subtype pre-trained segmentation model using all training datasets (Model 2), SDH model (Model 3), SAH model (Model 4), and small lesion (≤ 5 mL) model (Model 5). A summary of each model and the training dataset is shown in Figure 1. The DUMC and Qure.ai datasets were used for external testing.

**Figure 1.**
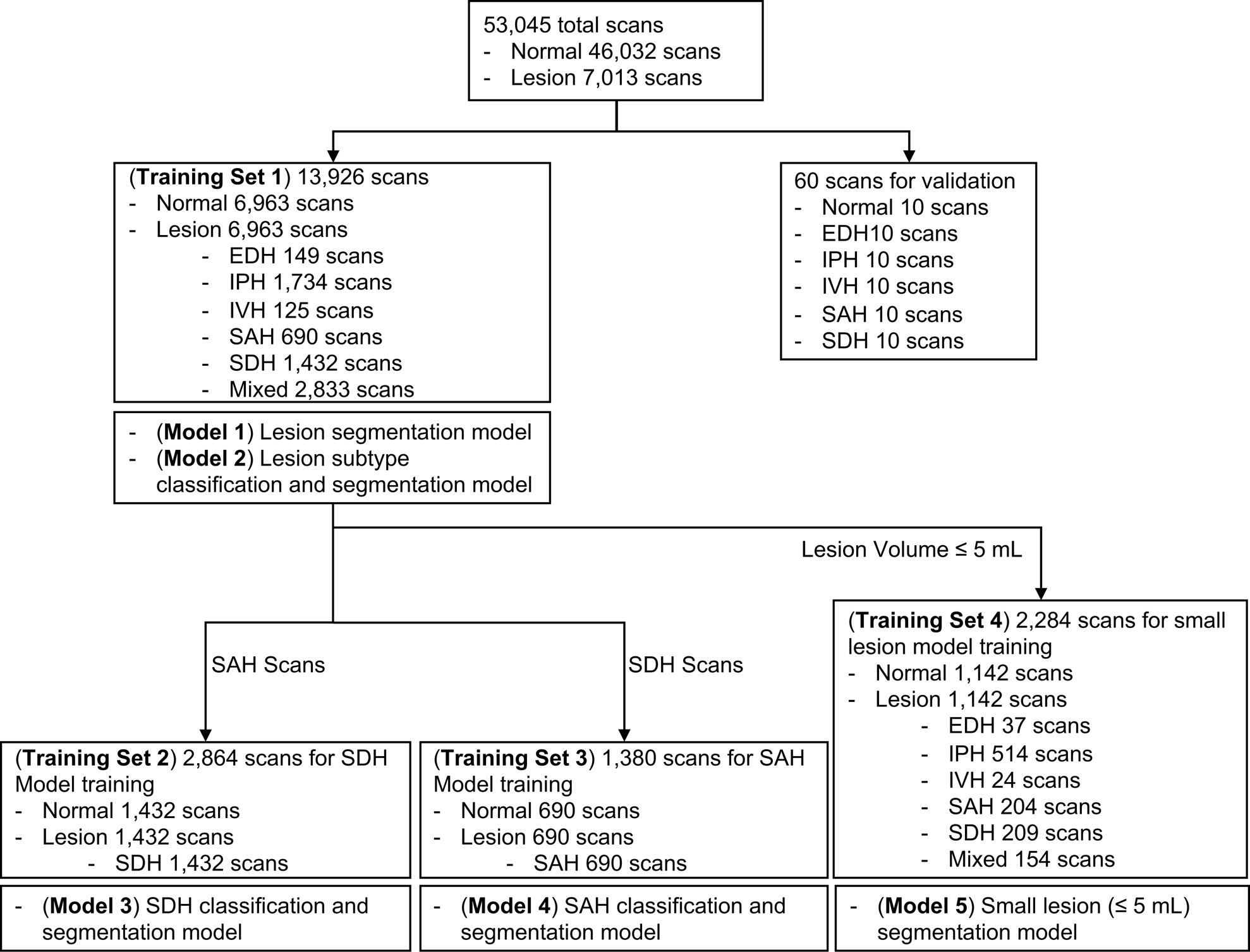
Summary of each deep-learning model and the training dataset used. EDH, epidural hemorrhage; IPH, intraparenchymal hemorrhage; IVH, intraventricular hemorrhage; SAH, subarachnoid hemorrhage; SDH, subdural hemorrhage.

#### 2.2. Ensemble base models

Five deep-learning models were trained using 2D U-net with the Inception module (eFigure 3).^15,16^ For the lesion subtype pre-trained segmentation model (Model 2), a pre-trained model in which down-sampling layers of U-net were pre-trained using hemorrhage subtype labeling was used. The Dice loss function, Adam optimizer, and a learning rate of 1e-4 were used for model training.

To determine whether the ensemble of base models (Models 1–5) improved the performance of hemorrhage detection in SAH, SDH, and small hemorrhage cases, we combined the base models and evaluated the performance of each combination model. From the two external test datasets, we extracted SAH, SDH, and small hemorrhage cases (with the same number of normal CT scans) to test the combination model.

#### 2.3. Weighted ensemble model

To ensemble the five base models using distinct datasets, their outputs needed to be assigned appropriate weight values according to the input data. Hence, we developed an additional weight model using input data comprising five-channel segmentation results from five base models, ranging from zero to one. Using random initiative weight values, the model was trained to select the weight values that minimized the Dice loss between the predicted segmentation at the pixel probability threshold of 0.5 and ground-truth segmentation (eMethods and eFigure 4 and 5).

### 3. Review of “difficult” ICH cases

After external testing of the weighted ensemble model, we defined the difficult ICH cases for expert reviews. “Difficult-for-AI” cases were chosen from the DUMC dataset when a) the probability of lesion ≤ 0.6 among cases annotated as hemorrhage or b) the probability of lesion ≥ 0.4 among cases annotated as no hemorrhage. “Difficult-for-humans” cases were selected from the Qure.ai dataset if the three annotators had not unanimously agreed with the ground truth during the initial labeling process.

Six neurology experts with six to eighteen years of clinical experience re-annotated the presence vs. absence of ICH in these two types of difficult images. The sensitivity, specificity, and accuracy of the weighted ensemble model and each expert were calculated. The inter-rater agreement among the experts was also calculated. If the ground truth and the opinions of six experts did not concur, a consensus meeting was held to amend the ground truth with a majority (4 or higher) vote. After the consensus meeting, the sensitivity, specificity, and accuracy of the weighted ensemble model and those of the six experts were re-calculated. Cases where the weighted ensemble model made incorrect predictions were subjected to qualitative assessments.

## Results

### Baseline characteristics of the datasets

The AI-Hub dataset, in which the presence of each hemorrhage subtype was labeled per slice and the hemorrhage was manually segmented, had 53,045 cases (mean 57.5 years, 47.5% female), and 13.2% (7,013) had ICH. In the RSNA dataset with the presence of each ICH subtype being labeled per slice, 39.6% (7,449) of 18,938 cases had ICH. Table 1 shows the proportion of each ICH subtype in the AI-Hub and RSNA datasets. The baseline characteristics of the external datasets from DUMC (n = 600) and Qure.ai (n = 386) are presented in Table 1.

**Table 1.**
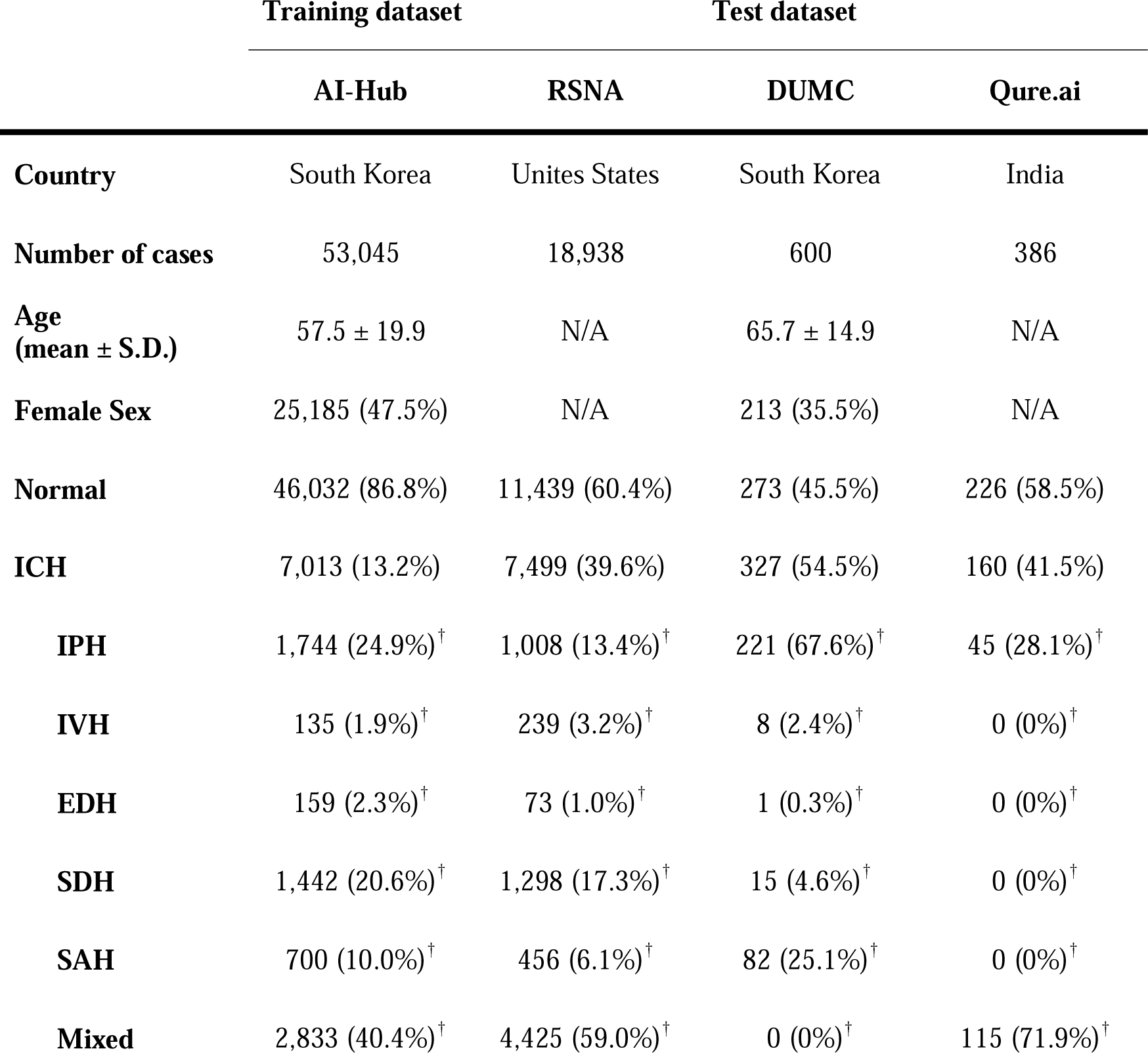
Baseline characteristics of the datasets. ^†^The percentages indicate the proportion of each subtype to the total number of lesions. RSNA, Radiological Society of North America; DUMC, Dongguk University Medical Center; ICH, intracranial hemorrhage; IPH, intraparenchymal hemorrhage; IVH, intraventricular hemorrhage; EDH, epidural hemorrhage; SDH, subdural hemorrhage; SAH, subarachnoid hemorrhage.

### Comparison of models trained with weakly vs. strongly annotated datasets

A dataset with strong annotations (AI-Hub dataset with location information) and two datasets with weak annotations (AI-Hub dataset without location information and RSNA dataset) were utilized for the training of four well-known deep-learning networks: DenseNet121, InceptionResNetV2, MobileNetV2, and VGG19. We tested four trained models on a composite of the DUMC and Qure.ai datasets. When trained using the RSNA dataset, the accuracies of DenseNet121, InceptionResNetV2, MobileNetV2, and VGG19 was 0.771 (95% C.I. 0.767–0.775), 0.770 (95% C.I. 0.766–0.774), 0.649 (95% C.I. 0.645–0.653), and 0.708 (95% C.I. 0.704–0.712), respectively. When trained using the AI-Hub dataset without location information, the accuracies were 0.812 (95% C.I. 0.809–0.816), 0.810 (95% C.I. 0.807–0.814), 0.645 (95% C.I. 0.641–0.650), and 0.707 (95% C.I. 0.705–0.711), respectively. When trained using the AI-Hub dataset with location information, the accuracies of all deep-learning networks except for DenseNet121 improved significantly, with the values being 0.756 (95% C.I. 0.753–0.761), 0.818 (95% C.I. 0.812–0.820), 0.658 (95% C.I. 0.655–0.664), and 0.862 (95% C.I. 0.859–0.865), respectively (Table 2 and eFigure 6).

**Table 2.** Comparison of models trained with weakly and strongly annotated datasets. Area under the curve (AUC), sensitivity, and specificity of four deep-learning networks trained on RSNA dataset, AI-Hub dataset without location information, and AI-Hub dataset with location information were shown. The AUC of each of the four deep-learning networks trained on the RSNA dataset was set as a reference, and the AUCs of the remaining models were compared using the DeLong test. C.I., confidence interval. *Standard error.

| 95% C.I. | RSNA dataset |  |  | AI-Hub without location dataset |  |  | AI-Hub with location dataset |  |  |
| --- | --- | --- | --- | --- | --- | --- | --- | --- | --- |
|  | AUC | Sensitivity | Specificity | AUC | Sensitivity | Specificity | AUC | Sensitivity | Specificity |
| **DenseNet121** | 0.771<br>(0.767 - 0.775) | 0.616<br>(0.611 - 0.620) | 0.754<br>(0.745 - 0.765) | 0.812<br>(0.809 - 0.816) | 0.554<br>(0.548 - 0.558) | 0.887<br>(0.880 - 0.895) | 0.756<br>(0.753 - 0.761) | 0.693<br>(0.688 - 0.698) | 0.657<br>(0.647 - 0.668) |
| P for AUC difference | Reference | | | $P = 0.0003$ (*0.0039) | | | $P < 0.0001$ (*0.0030) | | |
| **InceptionResNetV2** | 0.770<br>(0.766 - 0.774) | 0.623<br>(0.618 - 0.628) | 0.724<br>(0.716 - 0.736) | 0.810<br>(0.807 - 0.814) | 0.658<br>(0.654 - 0.663) | 0.825<br>(0.817 - 0.834) | 0.818<br>(0.812 - 0.820) | 0.745<br>(0.741 - 0.750) | 0.725<br>(0.715 - 0.735) |
| P for AUC difference | Reference | | | $P < 0.0001$ (*0.0031) | | | $P < 0.0001$ (*0.0031) | | |
| **MobileNetV2** | 0.649<br>(0.645 - 0.653) | 0.599<br>(0.595 - 0.605) | 0.600<br>(0.589 - 0.611) | 0.645<br>(0.641 - 0.65) | 0.599<br>(0.595 - 0.605) | 0.608<br>(0.598 - 0.620) | 0.658<br>(0.655 - 0.664) | 0.708<br>(0.704 - 0.713) | 0.499<br>(0.488 - 0.510) |
| P for AUC difference | Reference | | | $P = 0.4221$ (*0.0047) | | | $P = 0.0227$ (*0.0040) | | |
| **VGG19** | 0.708<br>(0.704 - 0.712) | 0.569<br>(0.564 - 0.574) | 0.754<br>(0.745 - 0.764) | 0.707<br>(0.705 - 0.711) | 0.460<br>(0.455 - 0.465) | 0.876<br>(0.869 - 0.884) | 0.862<br>(0.859 - 0.865) | 0.816<br>(0.812 - 0.820) | 0.731<br>(0.721 - 0.741) |
| P for AUC difference | Reference | | | $P = 0.6526$ (*0.0032) | | | $P < 0.0001$ (*0.0029) | | |

### Development of a weighted ensemble model

To improve the detection of SDH, SAH, and tiny lesions, which is regarded to be challenging, we designed a weighted ensemble model using multiple distinct datasets that not only had strong annotations but also reflected the various ICH features.

We first generated “Ensemble basic”, a weighted ensemble of two models, a lesion segmentation model (Model 1) and a subtype classification/segmentation model (Model 2), which were trained using CT images encompassing all ICH subtypes. Overall ICH detection accuracy measured by using a composite dataset of DUMC and Qure.ai, was 0.938 (95% C.I. 0.922–0.953, Table 3). The accuracy for SDH and SAH cases was respectively 0.893 (95% C.I. 0.865–0.919) and 0.944 (95% C.I. 0.942–0.945). Next, to further increase the accuracy in the diagnosis of SDH, SAH, and small lesions, we additionally developed models 3, 4, and 5, which were respectively trained on only SDH cases, only SAH cases, and only small lesions ≤ 5 mL. We then investigated whether the model performance was improved by combining Models 3, 4, or 5 with the weighted ensemble models for Models 1 and 2. “Ensemble SDH” showed an increased accuracy for SDH compared to “Ensemble basic”, from 0.893 (95% C.I. 0.865–0.919) to 0.927 (95% C.I. 0.903–0.948; *P* for AUC difference = 0.0002). For SAH, “Ensemble SAH” showed a comparable accuracy to “Ensemble basic” (0.952 [95% C.I. 0.951–0.954] vs. 0.944 [95% C.I. 0.942–0.945], *P* for AUC difference = 0.2439). “Ensemble small lesions” showed a comparable accuracy for total ICH compared to “Ensemble basic” (0.946 [95% C.I. 0.931–0.960] vs. 0.938 [95% C.I. 0.922–0.953], *P* for AUC difference = 0.1180). Finally, we developed a final weighted ensemble model that ensembles all models 1 to 5 and showed a significantly higher accuracy for total ICHs (0.951 [95% C.I. 0.937–0.964], *P* for AUC difference = 0.0379), compared to “Ensemble basic”.

**Table 3.**
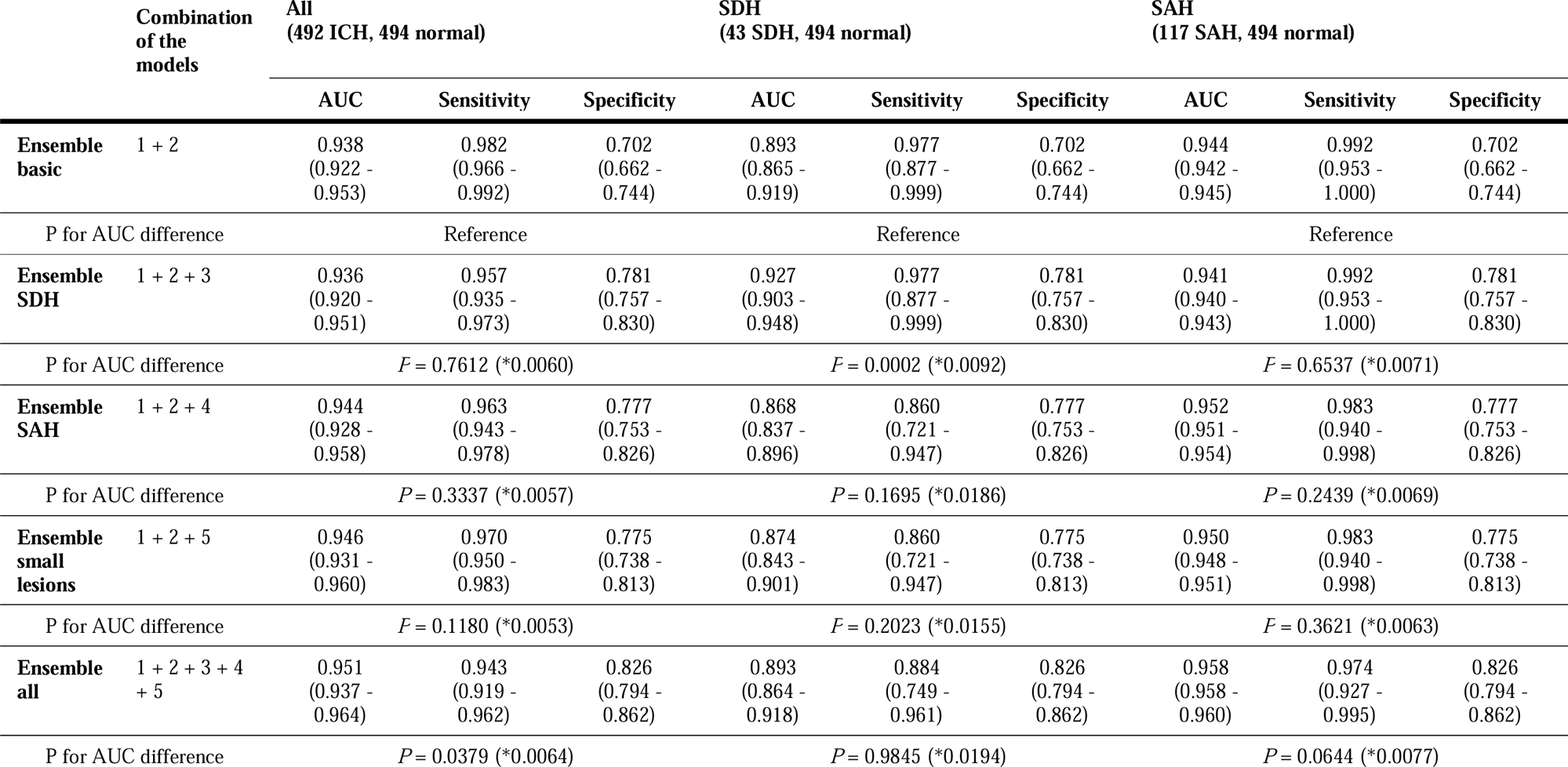
The structures of five weighted ensemble models and accuracies for all cases, SDH cases, and SAH cases. The area under the curves (AUCs) of “Ensemble basic” for total ICH, SDH, and SAH were set as references, and the AUCs of the remaining models were compared using the DeLong test. ICH, intracranial hemorrhage; SDH, subdural hemorrhage; SAH, subarachnoid hemorrhage. *Standard error.

### Comparison of the final weighted ensemble model with AI models that were previously built

We compared the performance of the final weighted ensemble model with that of DenseNet121, InceptionResNetV2, MobileNetV2, and VGG19 by using a test dataset that combines the DUMC and Qure.ai datasets. The weighted ensemble model significantly outperformed the other models in terms of sensitivity, specificity, and AUC (Figure 2 and eTable 1). Additional tests using either the DUMC or Qure.ai dataset showed similar results (eFigure 7).

**Figure 2.**
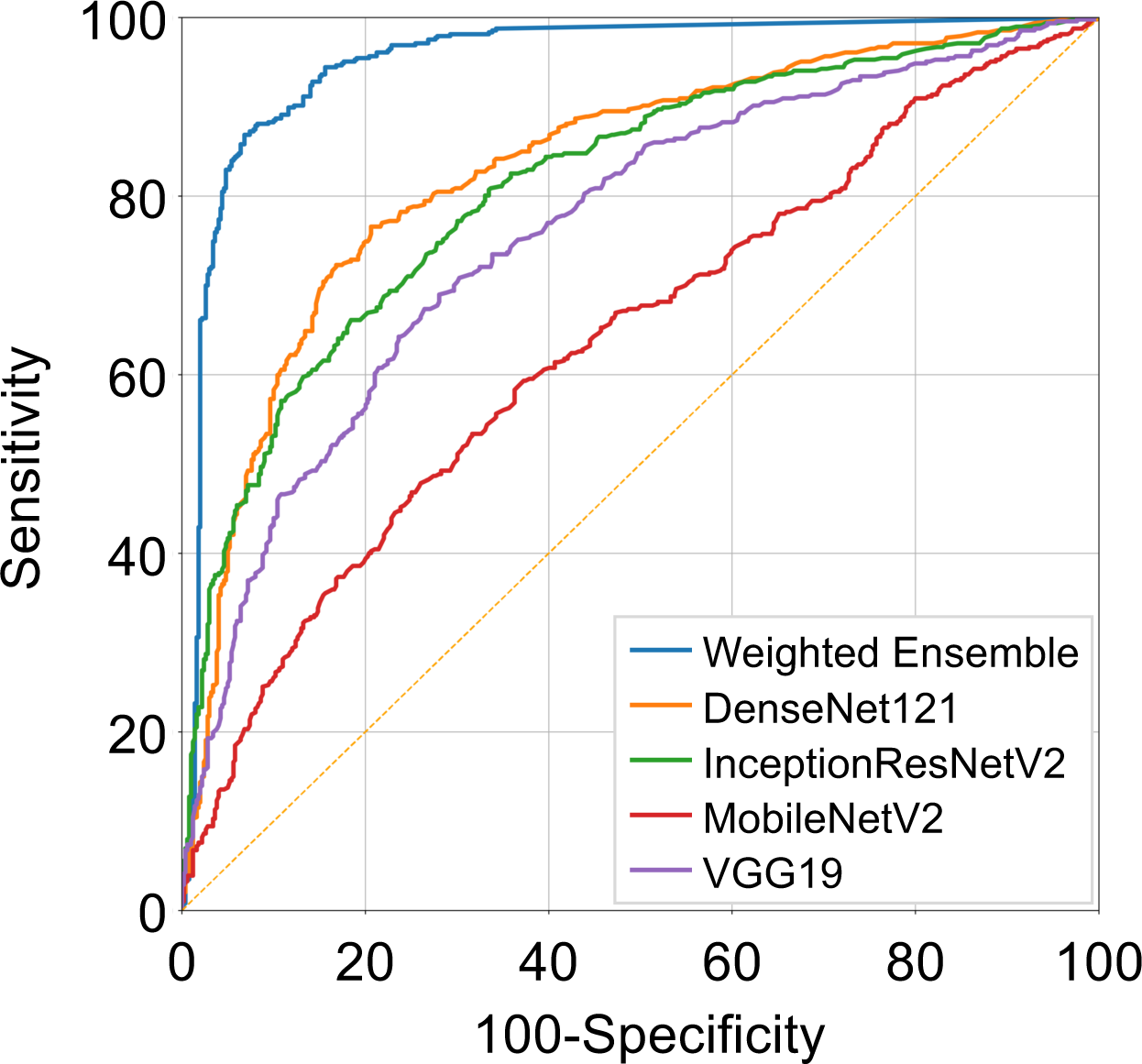
Receiver Operating Characteristic (ROC) curves representing the performance of the deep-learning models. DenseNet121, InceptionResNetV2, MobileNetV2, VGG19 trained on strongly annotated datasets and the final weighted ensemble model were applied to a test dataset combining the DUMC and Qure.ai datasets.

### Review of “difficult” 161 ICH cases

A total of 91 cases from the DUMC dataset were selected as difficult-for-AI cases: 17 cases from those classified as ICH that AI identified with a lesion probability of ≤ 0.6 and 74 cases from those classified as normal that AI identified with a lesion probability of ≥ 0.4 in external testing. A total of 70 cases from the Qure.ai datasets were selected as difficult-for-humans based on the three annotators’ disagreement. Six experts re-annotated these 161 cases for the presence vs. absence of ICH; there was complete agreement among the six experts for 81 cases, whereas there was at least one disagreement for 80 cases. For all 161 cases, the final weighted ensemble model showed an accuracy of 0.441, sensitivity of 0.431, and specificity of 0.462. The accuracies of the six experts were 0.671, 0.764, 0.708, 0.640, 0.683, and 0.714, with the interrater agreement (the Fleiss’ kappa value) being 0.536.

Among 95 cases where one or more experts disagreed with the initial ground truth, 80 were unanimous, five were disagreed upon by one expert, seven were disagreed upon by two experts, and three were disagreed upon by three experts. In 48 cases, the ground truth was changed following discussion and majority voting in the consensus meeting. The accuracy, sensitivity, and specificity of the weighted ensemble model for the revised ground truth was respectively 0.491, 0.457, and 0.525, without showing significantly differences when compared with those for the original ground truth. eTable 2 shows the qualitive assessments of the cases where the weighted ensemble model predicted incorrectly.

## Discussion

This study demonstrated that a) a deep-learning algorithm for detecting ICH trained with a strongly annotated dataset outperformed models trained with a weakly annotated dataset, and b) a weighted ensemble model that integrated separate models trained using SDH, SAH, or small-lesion (≤ 5 mL) ICH datasets achieved a higher AUC than four previous deep-learning models on external testing.

Medical image segmentation requires a lot of labor and resources.^17^ Although many transfer learning methods for weakly or partially annotated data have been developed,^18^ there is still a need for large-scale annotated data. To the best of our knowledge, no deep-learning algorithm for ICH detection has been developed using large-scale CT scans with segmentation annotation. We found that the accuracy of three of the four previously reported deep-learning models improved after training with strongly annotated datasets, compared to weakly annotated datasets.

Compared with magnetic resonance imaging, CT is less expensive, faster, and better at detecting ICH. However, some ICHs are more likely to be misdiagnosed due to their varied location and shape, small lesion size, and similar attenuation to adjacent tissue.^19^ For example, SDH an SAH are often difficult to distinguish from adjacent tissue,^4^ despite their distinct locations and shapes. Although there are classification methods for small lesions,^20^ a new deep learning strategy based on more comprehensive feature data may improve ICH detection performance. We achieved higher ICH detection accuracies using ensemble models that combined multiple separate models trained with datasets specialized for SDH, SAH, or small lesions. Employing the ensemble models, we also observed an increasing trend in specificity.

Despite recent development of many deep-learning algorithms for imaging diagnosis of ICH, their clinical application has yet to be accomplished. In addition to technical challenges such as the domain shift problem and the shortcut problem, there are also instances where determining the ground truth is difficult or inter-physician agreement is limited.^21–23^ After our expert meeting, as high as 30% (48/161) of difficult ICH cases, which however represented only 4.9% of the total test dataset cases (n = 986), required re-labeling of their ground truth. This may explain why it is challenging for AI to learn medical images that are difficult for experienced clinicians. Moreover, the re-labeling did not improve the accuracy of ICH detection by our weighted ensemble model trained with a total of 13,926 strongly annotated CT data. Further studies are required to investigate if fine-tuning a model after training with a larger high-quality training dataset, where difficult data are augmented, could increase the robustness, generalization, and discriminative power of the deep-learning algorithm.

Noisy labels have a negligible effect on the model performance in large datasets. In a handwritten number dataset, increasing the accuracy of random labels by only 1% significantly improved the model performance.^24,25^ However, if difficult cases are mixed at a low frequency across a dataset, noisy labels may affect ICH detection. There is a high demand in the medical profession for a model that can accurately diagnose both easy and complex cases. False positives and negatives may result in unnecessary and missed therapy, respectively. Future research should investigate if a) using ensemble models and b) expert review of difficult cases and adding them to a training dataset could overcome these challenges.

Our study has limitations. First, except for age and sex, no clinical information was available. Second, the classification and segmentation of some training data may not be accurate, because of the inclusion of difficult cases. Third, the proportions of mixed hemorrhages were high in the RSNA and AI-Hub datasets.

In conclusion, we developed a weighted ensemble model for ICH detection by training with strongly annotated CT scans obtained from multiple centers. Although challenging cases existed, external testing with a dataset from different ethnic origins demonstrated excellent performance of our model. We also showed that a better understanding and management of cases that are challenging for AI and humans is required to facilitate clinical use of ICH detection algorithms.

## Data Availability

All data produced in the present study are available upon reasonable request to the authors.

## Acknowledgment

This research was supported by the National Research Foundation of Korea and funded by the Ministry of Science and ICT (Grant NRF[2020M3E5D9079768). Wi-Sun Ryu had full access to all the data in the study and takes responsibility for the integrity of the data and the accuracy of the data analysis. Wi-Sun Ryu and Gi-Hun Park are employed by JLK Inc. Dong-Eog Kim is a stockholder of JLK Inc.

