## Supplementary Material for "Strengthening Deep-learning Models for Intracranial Hemorrhage Detection: Strongly Annotated Computed Tomography Images and Model Ensembles"

**eMethods.**

**eTable 1.** The performance of the weighted ensemble model and four well-known deep-learning networks, DenseNet121, InceptionResNetV2, MobileNetV2, and VGG19 on a test dataset.

**eTable 2.** The cases that the weighted ensemble model mispredicted among the “difficult” ICH cases.

**eFigure 1.** Four-channel input data for model development.

**eFigure 2.** Model development using weakly and strongly annotated ICH datasets.

**eFigure 3.** Schematic of the deep-learning model architecture used in this study.

**eFigure 4.** Comprehensive workflow of the developed weighted ensemble model.

**eFigure 5.** Schematic of weight model.

**eFigure 6.** Receiver Operating Characteristic (ROC) curves represent the performance of four well-known deep-learning networks according to the training datasets.

**eFigure 7.** Receiver Operating Characteristic (ROC) curves representing the performance of the deep-learning models for each test dataset.

**eMethods**

**1. Pre-processing**

A four-channel input image was used to develop the model. As previously described,^1-4^ CT windowing was used to generate three different images:1) a stroke window (width 40 and level 40), 2) brain window (width 80 and level 40), and 3) bone window (width 3,000 and level 500). A 3D U-net model developed in-house was used to strip the skull.^5^ We applied the skull stripping model to the 2) brain window image to create a 4) brain window with skull stripping (eFigure 1).

**2. Post-processing**

Five different hemorrhage detection models and the weighted ensemble model yielded five segmentation outputs and weight values per slice. The segmentation outputs and weight values were then multiplied using the following equation: The highest pixel probability value was selected as the maximal probability at the slice level (max[HemorrhageSlice]).

$$\boldsymbol{f(Slice)=max(}\frac{\sum_{\boldsymbol{i=1}}^{\boldsymbol{5}} \boldsymbol{SegOut}_{\boldsymbol{i}}\left( \boldsymbol{Pixel} \right)\boldsymbol{*}\boldsymbol{WeightOut}_{\boldsymbol{i}}\left( \boldsymbol{Pixel} \right)}{\boldsymbol{5}}\boldsymbol{)}$$

SegOut_i_ = output of each segmentation model; WeightOut_i_ = weight values for each segmentation model

We used the following equation to calculate the hemorrhage probability per case, in which max (HemorrhageSlice) indicates the maximal slice probability.

$$\boldsymbol{Probablity of Case=}\frac{\sum_{\boldsymbol{i=1}}^{\boldsymbol{number of slice with Hemorrhage}} \boldsymbol{HemorrhageSlice}_{\boldsymbol{i}}}{\boldsymbol{number of slice with Hemorrhage}}$$

**eTables.**

|  | **Total dataset** | | |
| --- | --- | --- | --- |
|  | **AUC** | **Sensitivity** | **Specificity** |
| **Ensemble model** | 0.953  (0.938 - 0.965) | 0.928  (0.901 - 0.949) | 0.857  (0.824 - 0.887) |
| P for AUC difference | Reference | | |
| **InceptionResNetV2** | 0.852  (0.828 - 0.873) | 0.802  (0.765 - 0.837) | 0.715  (0.674 - 0.755) |
| P for AUC difference | *P* < 0.0001 (*0.0128) | | |
| **DenseNet121** | 0.875  (0.852 - 0.895) | 0.856  (0.822 - 0.886) | 0.709  (0.667 - 0.749) |
| P for AUC difference | *P* < 0.0001 (*0.0119) | | |
| **VGG19** | 0.796  (0.770 - 0.821) | 0.751  (0.711 - 0.789) | 0.705  (0.663 - 0.745) |
| P for AUC difference | *P* < 0.0001 (*0.0144) | | |
| **MobileNetV2** | 0.650  (0.620 - 0.680) | 0.632  (0.588 - 0.675) | 0.573  (0.528 - 0.617) |
| P for AUC difference | *P* < 0.0001 (*0.0178) | | |

**eTable 1. The performance of the weighted ensemble model and four well-known deep-learning networks, DenseNet121, InceptionResNetV2, MobileNetV2, and VGG19 on a test dataset.** The external test datasets were a composite of the DUMC and Qure.ai datasets containing 487 cases with ICH and 499 cases without hemorrhage. All AUCs derived from other deep-learning models were significantly (all *P* < 0.001) lower than that of the Ensemble model. *Standard error.

| **False positive cases** |  | **False negative cases** |  |
| --- | --- | --- | --- |
| **Normal tissue** |  | SDH | 15 |
| Around falx | 10 | Hemorrhagic transformation | 7 |
| Transverse sinus | 6 | SAH | 4 |
| Bone artifact | 8 | Medulla hemorrhage | 1 |
| Vertebral artery | 2 | Thalamic hemorrhage | 1 |
| Medulla | 1 | BG IPH | 3 |
| Tentorium | 1 | Pontine hemorrhage | 1 |
| BG calcification | 2 | Contusional hemorrhage | 2 |
| **Abnormal tissue** |  | Other IPH | 3 |
| Metallic artifact | 3 | Tentorial hemorrhage | 1 |
| Motion artifact | 7 |  |  |
| skull defect-dura | 6 |  |  |
| Meningioma/tumor | 2 |  |  |
| **Total** | 48 | **Total** | 38 |

**eTable 2. The cases that the weighted ensemble model mispredicted among the “difficult” ICH cases.** BG, basal ganglia; SDH, subdural hematoma; SAH, subarachnoid hemorrhage; IPH, intraparenchymal hemorrhage.

**eFigures.**

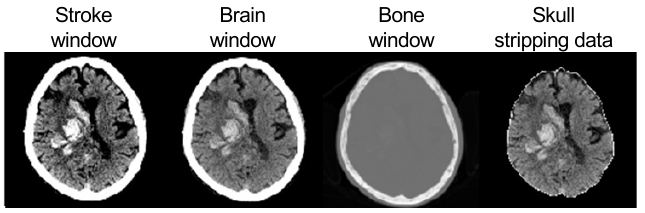

**eFigure 1. Four-channel input data for model development.** Stroke window (width 40 and level 40), brain window (width 80 and level 40), bone window (width 3,000 and level 500), and skull-stripped image.

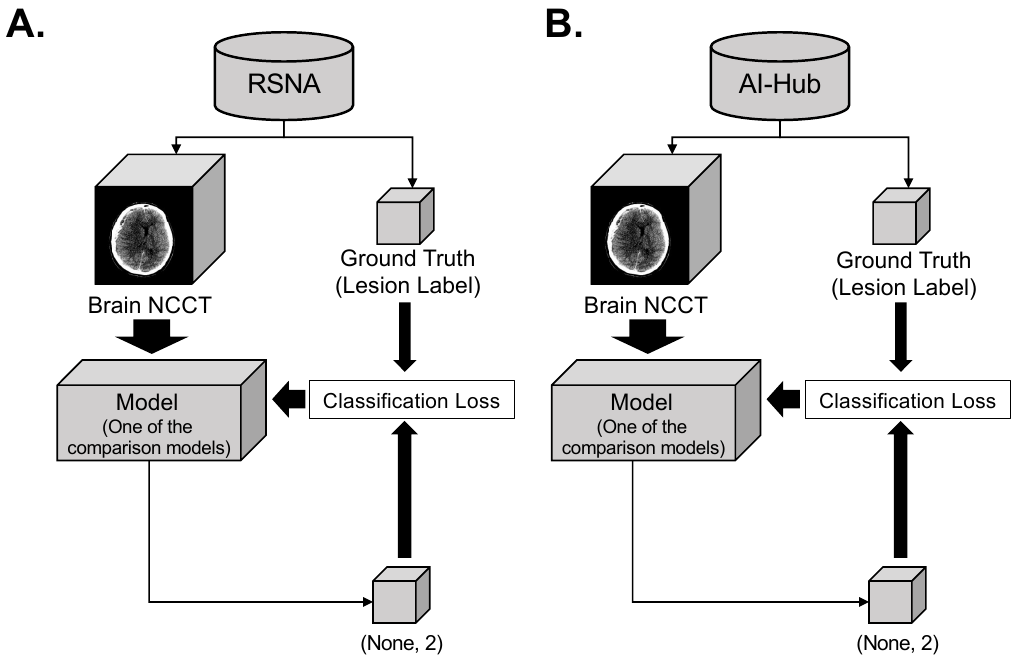

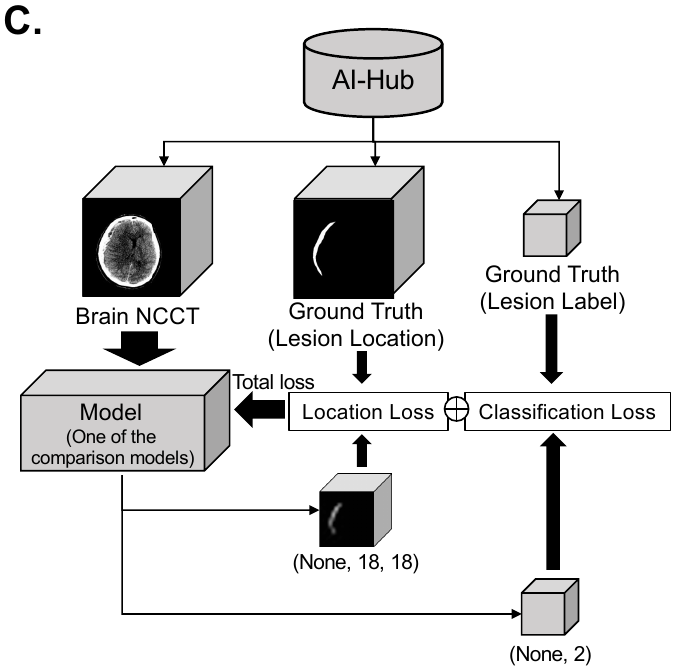

**eFigure 2. Model development using weakly and strongly annotated ICH datasets.** Training process using RSNA dataset (**A**), AI-Hub dataset without location (**B**), and AI-Hub dataset with location (**C**). ICH, intracranial hemorrhage; RSNA, Radiological Society of North America.

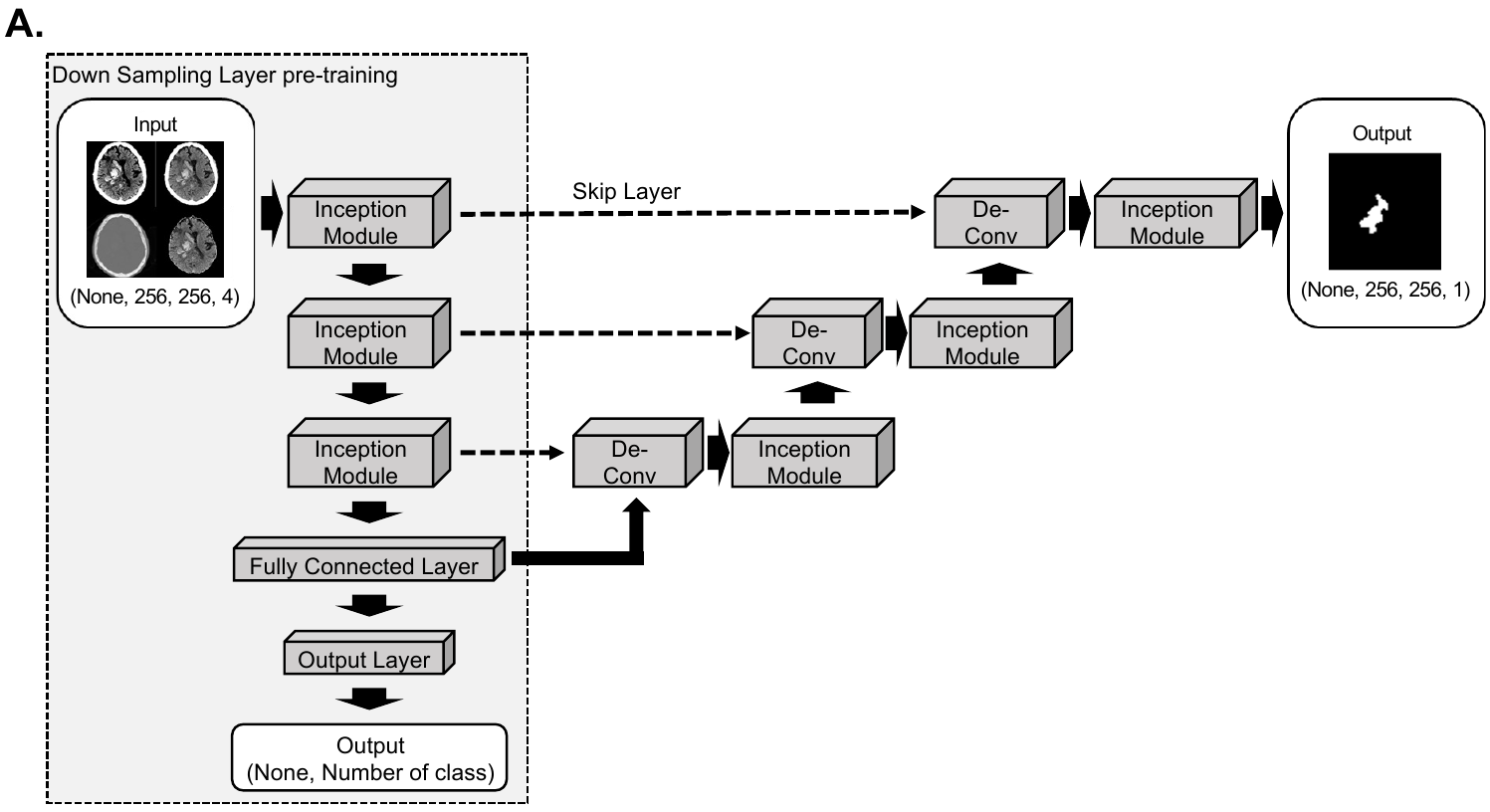

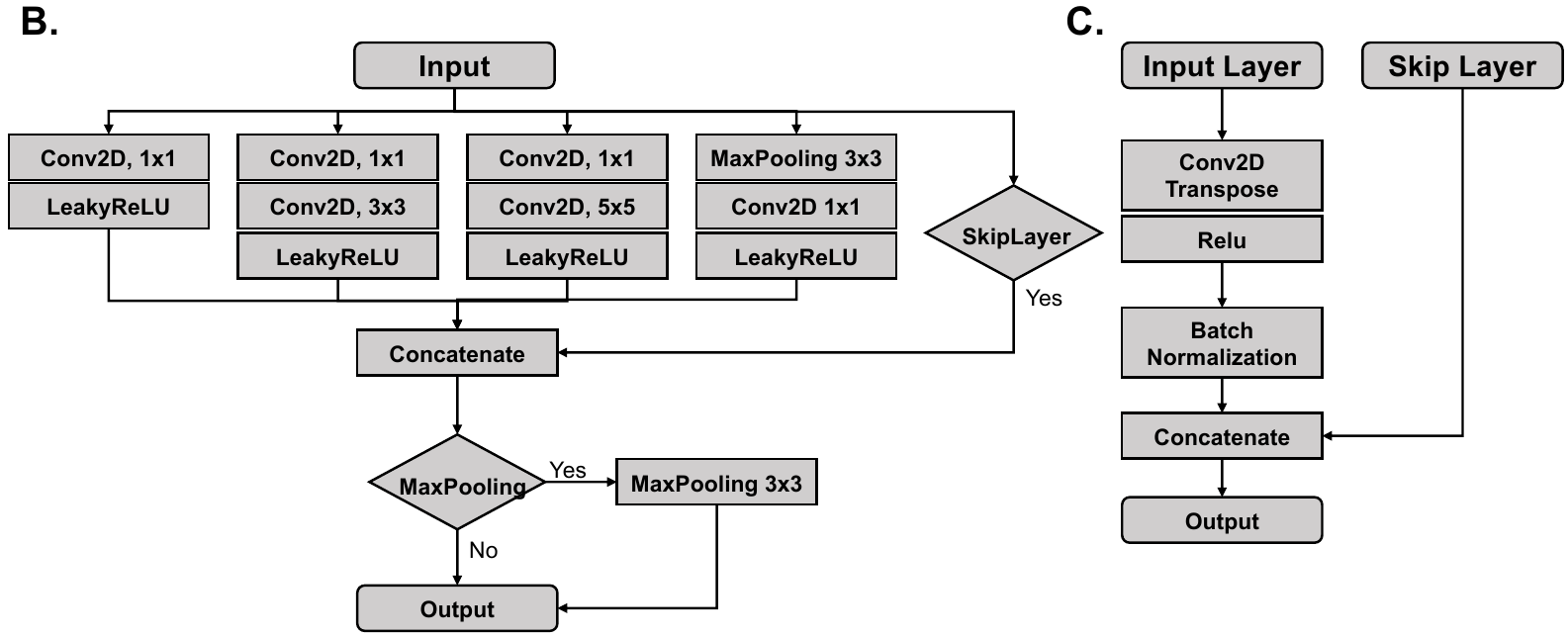

**eFigure 3. Schematic of the deep-learning model architecture used in this study.** **A.** Inception module-based U-Net architecture was used. **B.** Detailed architecture of the inception module. **C.** Detailed architecture of deconvolution block.

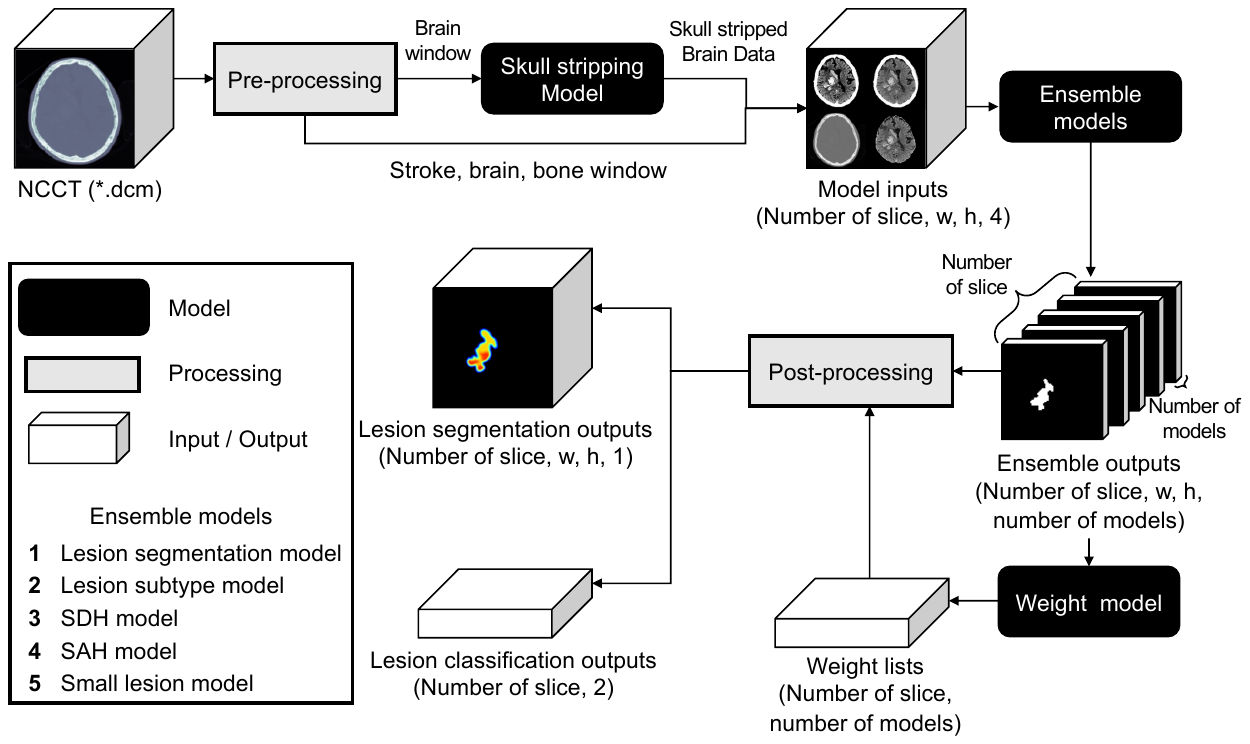

**eFigure 4.** **Comprehensive workflow of the developed weighted ensemble model.** The process includes pre-processing, skull stripping, ensembling models with weight model, and post-processing.

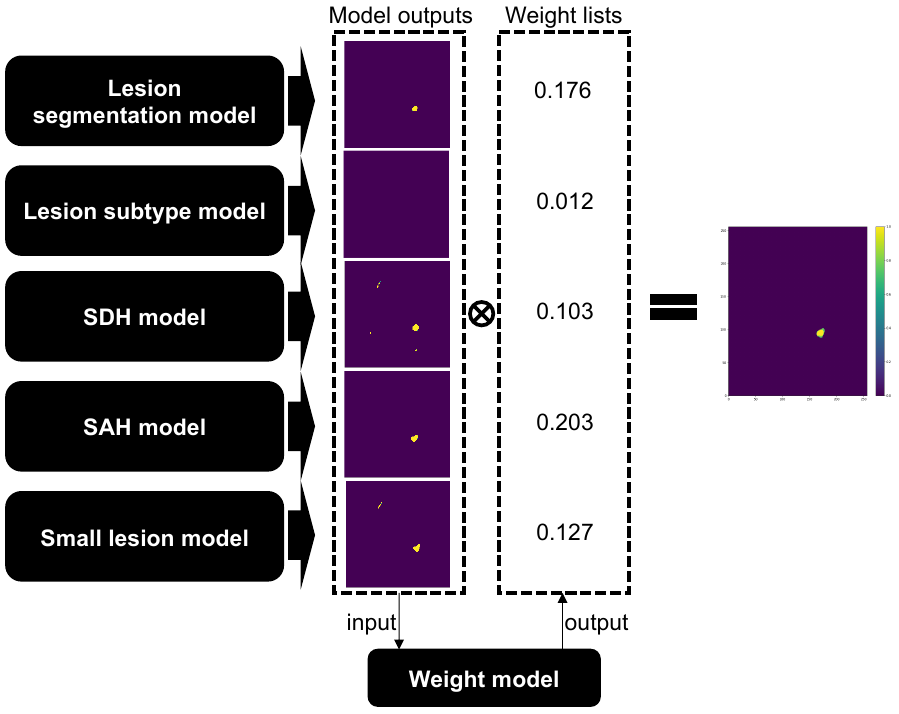

**eFigure 5. Schematic of weight model.** The model utilizes input data comprising the five-channel segmentation results obtained from the five base models. The final segmentation output is truncated at a threshold of 0.5. Each channel represents a probability value ranging from 0 to 1.

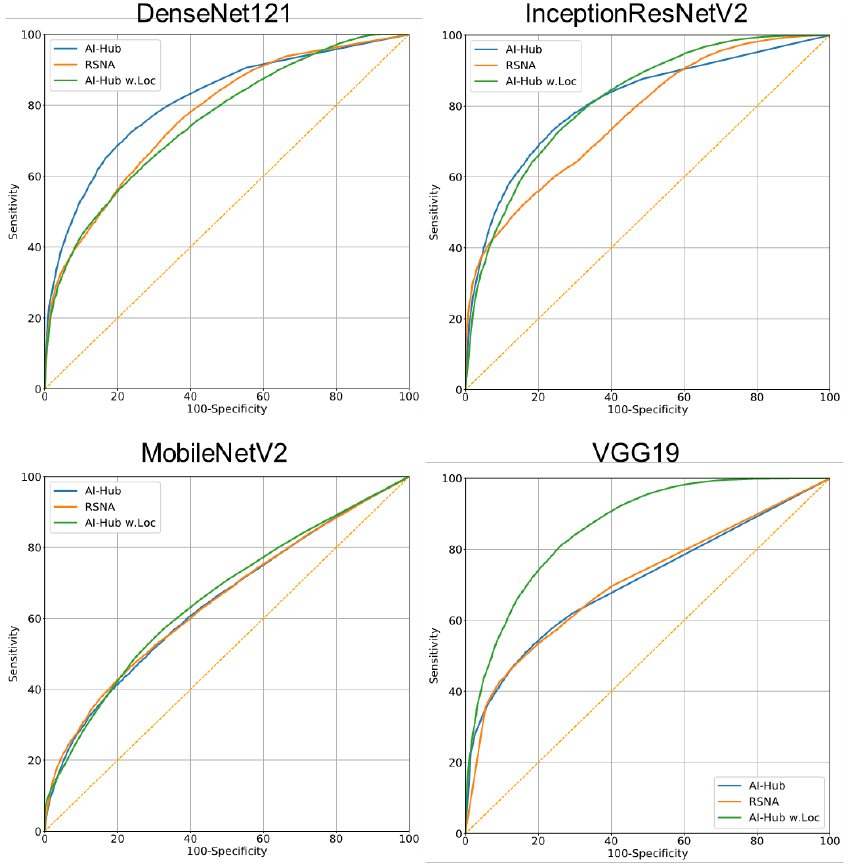

**eFigure 6. Receiver Operating Characteristic (ROC) curves represent the performance of four well-known deep-learning networks according to the training datasets.** The blue line indicates the AI-Hub dataset without location, the orange line indicates the RSNA dataset, and the green line indicates the AI-Hub dataset with location.

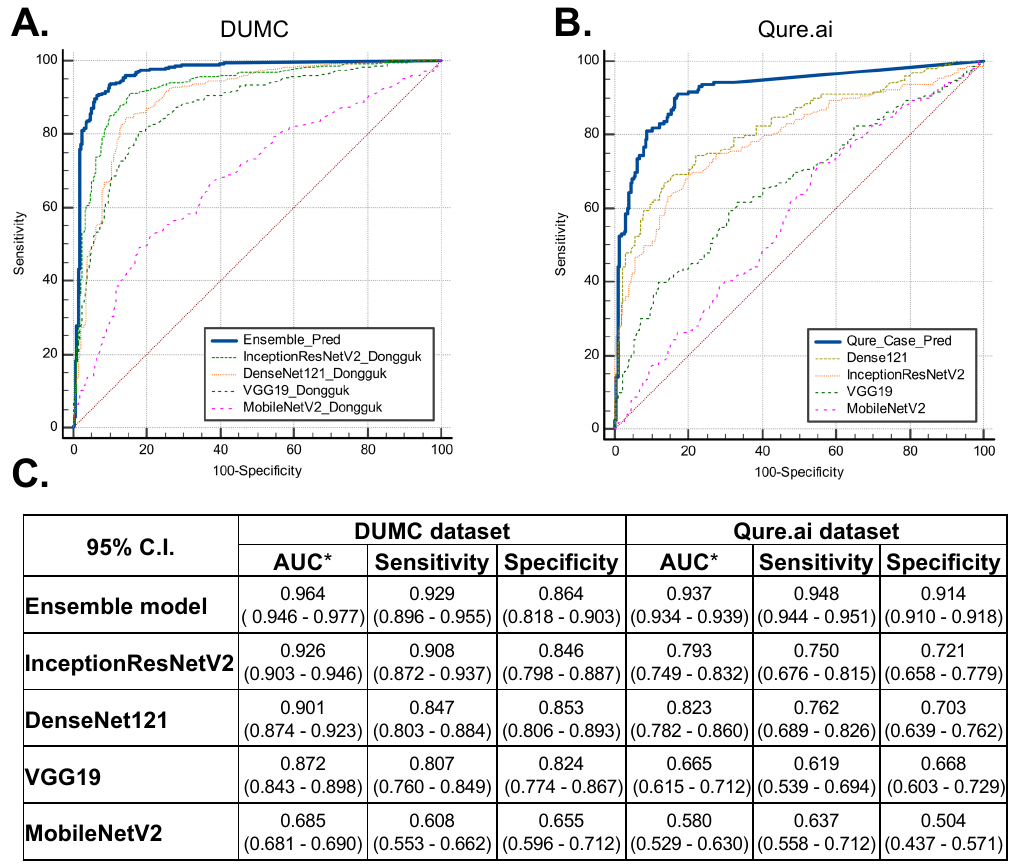

**eFigure 7.** **Receiver Operating Characteristic (ROC) curves representing the performance of the deep-learning models for each test dataset.** **A.** DenseNet121, InceptionResNetV2, MobileNetV2, VGG19 trained on strongly annotated datasets and the final weighted ensemble model were applied to the DUMC (**A**) and Qure.ai (**B**) dataset. **C.** AUC, sensitivity, and specificity of each deep-learning model on DUMC and Qure.ai datasets. DUMC, Dongguk University Medical Center; AUC, area under the curve. *All AUCs derived from the other deep-learning models were significantly (all *P* < 0.001) lower than those of the ensemble model.
